# Career Intentions of Final-Year Health-Professional Students in Lao PDR: A Cross-Sectional Study of Factors Influencing Public-Sector Attraction

**DOI:** 10.64898/2026.01.30.26345178

**Authors:** Sengthida Sivilay, Khampasong Theppanya, Bertrand Martinez-Aussel, Mick Soukavong, Mayfong Mayxay

## Abstract

**Background:** Ensuring a sufficient and motivated health workforce requires not only retaining existing staff but also understanding the intentions of those entering the labour market. In Lao PDR, limited civil-servant quotas, prolonged volunteer pathways, and expanding private sector opportunities shape the early career choices of health professional students. Yet little is known about how final-year students perceive the public sector or what influences their decision to join or avoid government service.

**Methods:** A cross-sectional survey was conducted among 298 final-year students from four major public health-training institutions. The questionnaire assessed demographic characteristics, motivations for choosing a field of study, post-graduation plans, and perceived drivers and barriers to public-sector employment. Descriptive statistics, chi-square tests, and multivariate logistic regression were used to identify factors independently associated with intention to work in the public sector.

**Results:** Results: Two thirds of students (66.1%) reported willingness to work in the public sector, though nearly as many simultaneously considered private sector employment (64.8%) and 43.3% expressed interest in working abroad, reflecting a “portfolio approach” to career planning under uncertainty. In multivariate analysis controlling age, field of study, and training institution, several factors independently predicted public sector intention. Each additional year of age increased the odds of public-sector preference by 21% (AOR 1.21, 95% CI 1.07-1.38, p = 0.003). Field of study demonstrated significant variation: pharmacy and dentistry students had 62% lower odds of public-sector intention compared to medical doctors (AOR 0.38, 95% CI 0.15-0.98, p = 0.045), while nursing and midwifery students showed equivalent preference (AOR 0.94, 95% CI 0.46-1.91, p = 0.855). Training institution emerged as a powerful predictor: students from provincial colleges demonstrated nearly three-fold higher odds of public-sector intention compared to those at the University of Health Sciences in Vientiane Capital (AOR 2.80, 95% CI 1.38-5.68, p = 0.004). Gender and marital status, while associated in bivariate analysis, did not remain significant in the adjusted model.

**Conclusion:** Final-year health professional students in Lao PDR demonstrate substantial public-sector commitment, but career intentions are shaped by institutional context and opportunity structures rather than motivation alone. To strengthen workforce recruitment, policymakers should leverage provincial training pipelines, implement field specific retention strategies for high risk disciplines, and ensure equitable career pathways that transform structural barriers into accessible entry mechanisms for all motivated graduates.

## 1. Introduction

A capable and motivated health workforce is central to achieving equitable population health outcomes and to realising Lao PDR’s Human Resources for Health Development Strategy 2021-2030 [1,2]. Although the country has expanded pre-service training over the past decade, the public sector continues to face systemic challenges including limited civil-servant quotas, persistent shortages of qualified personnel, and widening disparities across geographic areas. These structural constraints affect not only the retention of practising health workers but also the career intentions of students preparing to enter the labour market.

### Lao PDR Health Workforce Context

The health workforce landscape in Lao PDR is marked by both absolute shortages and pronounced maldistribution. According to the 24th HRH Technical Working Group presentation hold on December 2025, the national density of health workers stands at 3.35 per 1,000 population, falling to 2.85 per 1,000 when considering only civil servants. Levels well below international benchmarks for essential service coverage [3]. Urban centres report approximately 3.8 health workers per 1,000 compared with 1.4 per 1,000 in rural areas. Medical staff are even more unevenly distributed (1.6 vs. 0.4 per 1,000 population). More than 70% of health centres operate with fewer than five staff, and one-fifth with fewer than three. District hospitals frequently lack clinical specialists: Out of 148 only 56 have a family medicine specialist, 13 have a paediatrician and 10 have an obstetrician–gynaecologist [1,3].

Health professional education in Lao PDR has expanded over the past decade through a network of national and provincial public training institutions. Figure 1 shows their geographic distribution. Despite a steady increase in training output from the University of Health Sciences, the four Colleges of Health Sciences and five Schools of Public Health, the public sector’s absorption capacity remains constrained. In 2024, only 14.5% of newly created government staff quotas were allocated to the health sector, and less than 30% of the scholarships intended for HRH upgrading for 2024–2028 had been secured [2,3].

**Figure 1:**
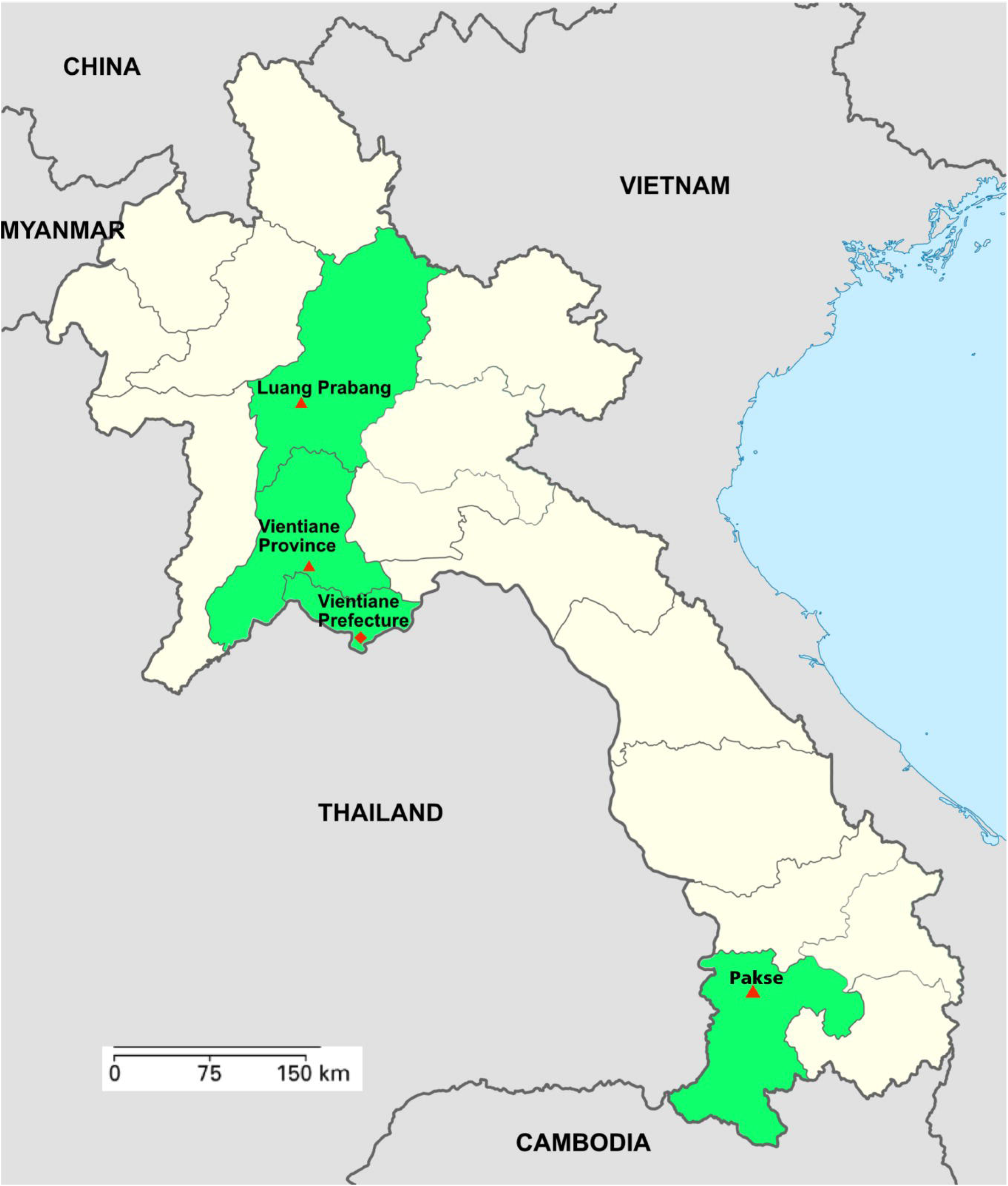

As a result, thousands of graduates depend on long-term volunteer positions, often for several years, with no guarantee of transition to a permanent post. For a considerable number of individuals, this period is characterised by financial hardship, constrained career progression and a pervasive sense of uncertainty. These factors have been identified as significant contributors to attrition among early-career professionals.

At the same time, the health labour market is changing. Growth in private hospitals and clinics in urban areas offers alternative employment pathways, often perceived as providing better remuneration or working conditions. The existence of regional migration pathways, specifically to Thailand, Japan and the Republic of Korea, has the effect of diversifying the career options available to young graduates. These shifts are of particular significance in shaping students expectations and influencing their perception of what constitutes a viable career.

The HRH Development Strategy 2021–2030 recognises these challenges and outlines a comprehensive reform agenda centred on strengthening HRH governance, meeting population needs, improving education quality and ensuring equitable distribution and retention. A cross-cutting emphasis is placed on stabilising the entry pipeline, including through more consistent recruitment processes, clearer incentive structures, and better forecasting of workforce supply and demand.

Yet, despite these policy commitments, little is known about how final-year students understand the opportunities and constraints ahead of them.

### Strategic Importance of Understanding Student Intentions

International studies show that students’ motivations, expectations and perceptions formed during training strongly influence whether they ultimately enter public service, choose private-sector employment, specialise, or migrate. In settings where the public sector cannot reliably offer early-career stability, students tend to diversify their options. This pattern has been documented in several Southeast Asian contexts but has not been empirically examined in Lao PDR in more than a decade [4,5].

Earlier national research on workforce retention in Lao PDR has focused primarily on practising health workers [6]. This work, together with national HRH assessments and policy discussions, has consistently identified quota scarcity, delayed remuneration and limited career progression as key challenges affecting public-sector retention. Similar structural constraints are already visible to students before graduation and may shape their career intentions at a critical transition point.

Despite the importance of these dynamics, Lao PDR lacks up-to-date evidence on how many students wish to enter the public sector, what motivates them to do so, and what deters them. Without such evidence, workforce planning risks misalignment between training output and real absorption capacity.

### Aim of this study

To our knowledge, this study addresses this gap by analysing the career intentions of final-year health-professional students across major public training institutions in Lao PDR. It quantifies their willingness to work in the public sector, identifies their motivations and concerns, and examines factors associated with public-sector preference. By providing timely evidence from the entry point of the workforce pipeline, the study supports national efforts to strengthen recruitment, rationalise quotas and ensure that investments in health education translate into a stable and competent health workforce.

## 2. Methods

### Study design and setting

A cross-sectional survey was conducted between May and August 2024 among final-year students from four major public health-training institutions: the University of Health Sciences (UHS) in Vientiane Capital, and the public health colleges of Luang Prabang, Vientiane Province, and Champasak. These institutions constitute the primary national pipeline for nurses, midwives, medical doctors, pharmacists and dentists.

### Sampling and participants

All final-year students in the selected institutions were invited to participate. Inclusion criteria were final-year status and consent to participate. The survey was self-administered electronically using mobile phones; students were gathered in designated sessions and provided with a secure access link, while a trained facilitator was present to respond to questions and provide clarification when needed. Participation was voluntary, anonymous, and based on informed consent provided via a checkbox on the first survey page.

### Survey instrument

The questionnaire captured:

- Demographics (age, gender, ethnicity, marital status, home province)
- Training information (institution, field of study)
- Reasons for choosing field of study (seven binary items)
- Post-graduation plans (public sector, private sector, volunteering, working abroad, leaving health sector)
- Motivations and deterrents for public-sector employment
- Reasons to prefer private-sector employment
- Survey usability feedback

Most variables were binary; attitudinal items used a five-point Likert scale. The instrument was delivered via KoboToolbox.

### Data collection

Data were collected between May and August 2024. No personal identifiers were collected. Data use was limited to research objectives, and participants could withdraw at any time without consequence. Data quality was verified daily by provincial coordinators who cross-checked completeness and logical consistency within KoboToolbox. Outliers and duplicate entries were reviewed manually before export to STATA version 16.0. Data were then cleaned using automated scripts, including missing-value flagging and range validation, to ensure analytic reliability.

A total of 298 valid responses were received, representing an overall response rate of 41%, with response rates ranging from 30% to 86% across institutions. Unique KoboToolbox logins prevented duplicate entries, and daily consistency checks ensured data integrity.

### Data analysis

All responses were exported to STATA version 16.0 for analysis. Descriptive statistics summarized sample characteristics and post-graduation intentions. Bivariate associations with intention to work in the public sector were examined using χ^2^ tests for categorical predictors and independent samples t-tests for continuous variables. Fisher’s exact test was employed where cell frequencies fell below five. Variables demonstrating associations at p < 0.20 in bivariate analysis were considered candidates for inclusion in multivariate models, following established guidelines for variable selection in logistic regression. Multivariable logistic regression was used to identify independent predictors. Results are presented as adjusted odds ratios (AOR) with 95% confidence intervals. Statistical significance was set at p < 0.05.

### Ethical considerations

The study protocol received approval from the Lao National Ethics Committee for Health Research (Approval No. 51/NECHR, 29 April 2024).

## 3. Results

A total of 298 final-year health-professional students completed the survey. The mean age was 23.2 years (range 18–47), and most respondents were women (72.5 %). The majority belonged to the Lao Loum ethnolinguistic group (81.5 %), and 91 % were single. Nursing students represented 57 % of the sample, followed by medical (19.8 %), midwifery (11.1 %), pharmacy (6.4 %) and dentistry (5.7 %). Just over one-third were enrolled at the University of Health Sciences (UHS) in Vientiane Capital, with the remainder studying at provincial public health colleges in Luang Prabang, Vientiane Province, or Champasak (Table 1).

### Reasons for choosing the field of study

Most students selected their field based on personal choice (81.5 %), although more than half reported having “no specific reason,” reflecting limited structured career guidance. Parental expectations (36.6 %), advice from relatives or acquaintances already working in the field (37.2 %), and practical considerations such as affordability (35.9 %) or school grades (34.6 %) also played an important role.

### Post-graduation intentions

Two-thirds of students (66.1 %) reported that they would consider working in the public sector after graduation, while a similar proportion (64.8 %) expressed interest in private-sector employment. One-third (32.9 %) stated that they would accept a volunteer position to secure a fixed post in the government system. Working abroad was considered by 43.3 % of respondents, whereas only 2 % indicated a desire to work outside the health sector (Table 2).

### Motivations and deterrents

Students attracted to the public sector primarily cited stability and job security (55.0 %), followed by professional development opportunities (30.2 %), contribution to national development (25.5 %), and community service motivations (24.2 %). Conversely, the most common deterrents were structural: lack of civil-servant quotas (21.1 %) and reluctance to volunteer for long periods (19.5 %). Low salary was cited by only 3.4 %.

Reasons to prefer private-sector work centred on financial and professional advantages: better incomes (48.3 %), career opportunities (27.9 %), bonuses (26.2 %), and more favourable working environments (21.1 %).

### Determinants of Intention to Work in the Public Sector

Multivariate logistic regression analysis (Table 3) identified several independent predictors of public-sector employment intention among final-year health professional students in Lao PDR. The final adjusted model included age, field of study, and training institution as significant predictors, while gender, marital status, and ethnicity were not retained in the final model due to non-significant associations in adjusted analysis. Age demonstrated a significant positive association with public-sector intention. Each additional year of age was associated with a 21% increase in the odds of intending to work in the public sector (AOR 1.21, 95% CI 1.07–1.38, p = 0.003). While the crude odds ratio was more modest (OR 1.09), adjustment for field of study and training institution strengthened this relationship.

Substantial variation in public-sector intention emerged across health professional disciplines. Using medical doctor students as the reference category (72.88% expressing public-sector intention), pharmacy and dentistry students demonstrated markedly lower public-sector preference. Only 38.89% of pharmacy/dentistry students intended to work in the public sector a striking 34 percentage point difference from medical doctors. In adjusted analysis, pharmacy and dentistry students had 62% lower odds of public-sector intention compared to medical doctors (AOR 0.38, 95% CI 0.15–0.98, p = 0.045). In contrast, nursing and midwifery students showed no significant difference from medical doctors in their public-sector preferences. With 68.97% planning to enter government service, nursing/midwifery students demonstrated statistically equivalent public-sector intention to medical doctors (AOR 0.94, 95% CI 0.46–1.91, p = 0.855). This convergence suggests relatively stable public-sector orientation among nursing and midwifery cadres, who represent the numerical majority of health professional students.

Training institution emerged as a highly significant predictor of career sector preference. Students from provincial health training institutions demonstrated substantially higher public-sector intention (72.04%) compared to those enrolled at the University of Health Sciences (UHS) in Vientiane Capital (56.25%) a difference of nearly 16 percentage points. The crude odds ratio suggested provincial students were twice as likely to prefer public-sector careers (OR 2.00), but this relationship strengthened considerably in adjusted analysis (AOR 2.80, 95% CI 1.38–5.68, p = 0.004).

## 4. Discussion

This study provides an unusual window into the expectations and hesitations of final-year health-professional students in Lao PDR at the exact moment they prepare to enter a labour market defined by quota scarcity, economic uncertainty and expanding private-sector opportunities. What emerges is not a lack of interest in serving the public, but rather a portrait of students navigating a system where the path into government service has become increasingly narrow.

Roughly two-thirds of students expressed willingness to work in the public sector. This proportion is encouraging and broadly consistent with findings from other low- and middle-income settings, where public-sector commitment often remains high prior to graduation. Studies in India and China report that soon-to-be graduates frequently value ideals of service, stability, and community contribution, even as they anticipate better salaries or working conditions elsewhere [7,8]. Yet, similar to patterns observed in several Southeast Asian and South Asian settings, our respondents keep multiple options open, simultaneously considering public-sector employment, private-sector work and migration [4,9]. This reflects what medical-education researchers have called the “portfolio approach” to early career planning: when uncertainty is high, students diversify rather than commit.

The factors shaping these intentions in Lao PDR show both convergence with international experience and distinct local dynamics. The strongest predictor of intending to work in the public sector, the willingness to volunteer, is deeply context-specific. In most countries, early career choices hinge on salary expectations, supervision, or family circumstances. In Lao PDR, however, the volunteer system has become the de facto gatekeeper to public employment. Students who accept this reality, or feel they have no alternative to it, are far more likely to imagine themselves in government service. Those who reject years of unpaid or low-paid volunteering understandably gravitate toward private or international routes. This dual meaning of volunteering, as both a stepping stone and a deterrent, has been noted informally in workforce studies but rarely quantified. Our findings document it clearly.

The training institution also exerts a significant influence on the outcome. It was found that students who had undergone training at the University of Health Sciences and had been exposed to urban private-sector facilities and more competitive employment norms were significantly less inclined to pursue a career in the public sector. Similar patterns have been observed in Thailand and the Philippines, where urban training environments are associated with higher salary expectations and stronger private-sector pull [9,10]. Conversely, students trained in provincial institutions in Lao PDR resemble findings from rural-origin students in Thailand’s CPIRD/ODOD tracks: those who study closer to home or in smaller settings are more likely to envision themselves serving in government facilities [11].

Economic vulnerability also shapes choices. Students who selected their field because it was the most affordable training option were more likely to prefer the public sector. This echoes evidence from India and Kenya, as well as other low- and middle-income settings, where students and early-career health workers from more modest socio-economic backgrounds often prioritise job stability over salary maximisation [7,12]. Interestingly, very few respondents explicitly cited low salary as a reason to avoid the public sector. This does not suggest that salary is irrelevant, but rather that many have not yet confronted the realities of late payments or the rising cost of living issues that strongly affect practising health workers. Their expectations may be optimistic now, but vulnerable to rapid disillusionment if conditions remain unchanged.

The intention to work abroad, while common, did not independently reduce public-sector intention after controlling for other variables. This finding mirrors evidence from Vietnam and South Asian settings, where migration aspirations among students and early-career health workers often represent longer-term or contingent possibilities rather than immediate plans. Many foresee working locally first to gain experience before considering international opportunities at a later stage [4,13].

Overall, the portrait that emerges is one of students who are motivated, adaptable, and generally well disposed toward public service, but whose career decisions will ultimately be shaped by the structure of opportunity, not the absence of commitment.

In addition to the quantitative findings presented here, students also provided open-ended suggestions regarding training and career pathways, which are summarised in the Supplementary Material.

### Policy implications

The findings of this study highlight a limited number of policy levers that could substantially improve attraction to the public sector and strengthen the transition from training to employment. Evidence from comparable settings suggests that these levers are most effective when applied at the entry point of the health workforce pipeline. Table 4 summarises these recommendations

#### a) Structuring the volunteer pathway as a formal entry stage

Volunteering currently constitutes an implicit entry route into public-sector employment. Rather than functioning as an informal filter, this pathway could be formalized into a regulated early-career stage, as observed in several neighboring countries.

In Thailand and Vietnam, early-career public-sector service is generally organised through formal arrangements that define duration, remuneration, supervision, and pathways to longer-term employment [10,14]. Drawing on these experiences, introducing maximum durations for volunteer service, basic stipends or allowances, and transparent criteria for transition to contractual or civil-servant positions would improve predictability and equity. Prioritizing long-serving volunteers in quota allocation would further reinforce institutional trust and reduce early attrition.

#### b) Improving transparency of recruitment quotas

Uncertainty surrounding civil-servant recruitment undermines confidence in public-sector careers. Experience from Indonesia and other comparable settings suggests that, even when recruitment capacity is limited, regular and transparent communication of quotas improves alignment between graduate expectations and system capacity [15].

Routine dissemination of expected national quotas, provincial allocation, qualification requirements, and priority cadres would allow students and training institutions to better anticipate opportunities and plan accordingly.

#### c) Strengthening career guidance during pre-service training

Embedding structured career guidance during pre-service education would support more informed decision-making. International evidence indicates that early exposure to public-sector practice combined with clear information on recruitment mechanisms is feasible and acceptable to stakeholders [16]. Experience from Thailand’s CPIRD and ODOD programs further demonstrates that such approaches can increase uptake of public and rural posts [10,11]. Integrating career counselling, briefings on quota processes and scholarship obligations, and structured rural or district placements into training curricula would help align student expectations with labor-market realities.

#### d) Reducing disparities between central and provincial training pathways

Differences in public-sector preference between centrally and provincially trained students mirror patterns observed in other countries. Targeted measures such as bonded public-sector scholarships, incentivised rural rotations, and structured early-career placements have been used in Thailand and the Philippines to counterbalance urban private-sector pull [9,10,11]. Applying similar mechanisms in Lao PDR could help maintain public-sector attractiveness among graduates trained in central institutions.

#### e) Supporting financial security at workforce entry

Measures that enhance financial predictability at the start of public-sector careers have proven effective in multiple low- and middle-income countries. Interventions such as timely salary payment, housing support, and access to low-interest loans, implemented for example in Rwanda and Nepal, have improved early-career retention without requiring major wage reforms [17,18].

Ensuring basic financial security for new public-sector entrants would strengthen retention among graduates entering through volunteer or contractual arrangements.

#### f) Using student-intention data for workforce planning

Finally, institutionalising the analysis of student career intentions would support more responsive HRH planning. Evidence from Malaysia and comparable settings shows that such data can inform workforce forecasting and help align training output with recruitment and deployment capacity [19].

In Lao PDR, regular use of student-intention data could inform training intake, provincial planning and long-term projections, reducing inefficiencies and avoidable loss of motivated graduates.

### Study Limitations

The survey sample is broadly representative of final-year students in major public health-training institutions. However, several limitations should be considered when interpreting these findings.

First, the data are based on self-reported intentions rather than observed employment outcomes. What students say at graduation may not reflect what they ultimately do once confronted with the realities of income, family needs or available positions. As seen in international longitudinal studies, early optimism can give way to rapid career drift.

Second, the analysis is cross-sectional, capturing a single moment in time during a period of economic difficulty in Lao PDR. Intentions are likely sensitive to macroeconomic fluctuations, policy announcements and local labor-market conditions.

Third, the logistic regression model identifies associations, not causation. Some predictors such as willingness to volunteer or preference for private-sector work may partially reflect underlying personality traits, expectations or economic constraints that were not directly measured.

Fourth, several variables (e.g., reasons to work in the public sector) were only answered by students already expressing public-sector willingness, limiting comparative analysis. Broader attitudinal measures would allow more nuanced modelling.

Fifth, the sample includes four major training institutions but does not cover all schools in the country. The findings remain highly relevant nationally, but some subgroups particularly private-school students are not represented.

Finally, although the dataset is complete and internally consistent, minor reporting biases cannot be excluded. Students may have responded in ways they believed socially appropriate, particularly regarding public-sector motivation.

Despite these limitations, the study fills a critical evidence gap in Lao HRH planning. It provides a rare, empirical insight into the future workforce pipeline at a moment when public-sector absorption is shrinking and competition from private and international markets is growing.

## 5. Conclusion

This study offers a rare look at the hopes and constraints shaping the career intentions of final-year health-professional students in Lao PDR. The results show that most students remain genuinely open to public-sector service, motivated by stability, community impact, and the promise of professional development. However, this willingness is accompanied by a realistic awareness of the structural barriers that have come to define early employment in the health system. The most notable of these barriers are quota shortages, prolonged volunteer periods, and the perceived competitiveness of private-sector opportunities.

Intentions are not shaped by lack of commitment, but by the architecture of opportunity. Students who are ready or able to navigate the volunteer pathway are dramatically more likely to imagine a future in government service, while those facing tighter economic constraints or exposed to more attractive alternatives often look elsewhere. The divergence between UHS students and those from provincial institutions underscores how training environments condition expectations and career trajectories.

These findings have direct implications for the Human Resources for Health Development Strategy 2021–2030 [2]. Strengthening the public-sector pipeline will require deliberate action: formalizing the volunteer system into a fair and time-bound entry mechanism, enhancing transparency around quotas, supporting early-career financial security, and ensuring that students receive realistic and supportive career guidance throughout their training.

Ultimately, Lao PDR has no shortage of motivated future health workers. What is missing is a system that reliably transforms their motivation into sustainable public-sector careers. By addressing the barriers at the very start of the professional journey, the Ministry of Health can safeguard its future workforce and advance national goals of equitable, competent and resilient health services.

## Data Availability

All data produced in the present study are available upon reasonable request to the authors

## Declarations

### Ethics approval and consent to participate

Ethical approval for this study was granted by the Lao National Ethics Committee for Health Research (Approval No. 51/NECHR, dated 29 April 2024) under the research proposal titled *“Understanding the Factors Affecting the Attraction and Retention of Health Professionals in Lao PDR*.*”* All participants provided informed consent. Participation was voluntary, anonymous, and conducted in accordance with national ethical guidelines and international standards.

### Consent for publication

Not applicable.

### Availability of data and materials

The datasets generated and analysed during the current study are not publicly available due to privacy and confidentiality agreements but may be obtained from the corresponding author upon reasonable request.

### AI assistance

During the preparation of this manuscript, the authors used an artificial intelligence tool (ChatGPT, OpenAI) to support language editing, structural refinement, and clarity enhancements. All research design, data analysis, interpretation, and final manuscript approval were conducted exclusively by the human authors.

### Authors’ contributions

- **Sengthida Sivilay**: Led data collection, performed preliminary analysis, and drafted the initial manuscript.
- **Khampasong Theppanya**: Supported field coordination, contributed to data validation, and reviewed key findings.
- **Bertrand Martinez-Aussel**: Provided methodological guidance, assisted in data interpretation, and led substantive revisions.
- **Mick Soukavong**: Conducted statistical analysis and contributed to the interpretation of results.
- **Mayfong Mayxay**: Conceived and initiated the study, supervised research implementation, and critically reviewed and approved the final manuscript.

All authors read and approved the final version of the manuscript.

## Acknowledgements

The authors would like to express their sincere gratitude to the participating students and the human resources and administrative departments of the educational institutions that facilitated this study. Special thanks are extended to the World Health Organization (WHO WPRO and Lao Country Office) for their generous financial and technical support.

