## Supplementary material for "Career Intentions of Final-Year Health-Professional Students in Lao PDR: A Cross-Sectional Study of Factors Influencing Public-Sector Attraction": Tables

**Table 1.** Characteristics of final-year health-professional students (n = 298)

| **Characteristic** | **n** | **%** |
| --- | --- | --- |
| **Age (years)** |  |  |
| Mean (SD) | 23.2 (±3.7) | – |
| Median (range) | 22 (18–47) | – |
| **Gender** |  |  |
| Female | 216 | 72.5 |
| Male | 78 | 26.2 |
| Prefer not to say | 4 | 1.3 |
| **Ethnolinguistic group** |  |  |
| Lao Loum | 243 | 81.5 |
| Lao Soung | 33 | 11.1 |
| Lao Theung | 20 | 6.7 |
| Tai Dam/Deng | 2 | 0.7 |
| **Marital status** |  |  |
| Single | 271 | 91.0 |
| Married | 24 | 8.1 |
| Divorced/Separated | 2 | 0.7 |
| Widow | 1 | 0.3 |
| **Training institution** |  |  |
| UHS Vientiane Capital | 112 | 37.6 |
| Luang Prabang PH College | 79 | 26.5 |
| Vientiane Province PH School | 54 | 18.1 |
| Champasak PH College | 53 | 17.8 |
| **Field of study** |  |  |
| Nursing | 170 | 57.0 |
| Medicine | 59 | 19.8 |
| Midwifery | 33 | 11.1 |
| Pharmacy | 19 | 6.4 |
| Dentistry | 17 | 5.7 |

**Table 2:** Post-graduation intentions and motivations (n = 298)

| **A. Plans after graduation** | **n** | **%** |
| --- | --- | --- |
| Intend to work in the public sector | 197 | 66.1 |
| Intend to work in the private sector | 193 | 64.8 |
| Willing to volunteer for a permanent post | 98 | 32.9 |
| Intend to work abroad | 129 | 43.3 |
| Intend to work outside health sector | 6 | 2.0 |
| **B. Reasons to work in the public sector**  *(Multiple responses allowed; denominators = 298)* | **n** | **%** |
| Stability and job security | 164 | 55.0 |
| Professional development & career advancement | 90 | 30.2 |
| Contribution to national development | 76 | 25.5 |
| Service to community / sense of purpose | 72 | 24.2 |
| Government benefits | 43 | 14.4 |
| Follow relatives/friends | 25 | 8.4 |
| **C. Reasons *not* to work in the public sector** | **n** | **%** |
| No available civil-servant quota | 63 | 21.1 |
| Do not want to volunteer too long | 58 | 19.5 |
| Better private-sector opportunities | 30 | 10.1 |
| Prefer to work abroad | 14 | 4.7 |
| Do not want remote postings | 14 | 4.7 |
| Do not want to leave family | 13 | 4.4 |
| Low salary | 10 | 3.4 |
| **D. Reasons to work in the private sector** | **n** | **%** |
| Better incomes | 144 | 48.3 |
| Better career opportunities | 83 | 27.9 |
| Better bonuses | 78 | 26.2 |
| Better working conditions | 63 | 21.1 |
| Better management | 55 | 18.5 |
| Open-ended contracts | 37 | 12.4 |

**Table 3:** Crude and adjusted odds ratios of factors associated with intention to work in the public sector

| Variable | Intend to work in public sector (%) | Crude OR | Adjusted OR | 95% CI | p-value |
| --- | --- | --- | --- | --- | --- |
| Age | — | 1.09 | 1.21 | 1.07–1.38 | 0.003 |
| Gender |  |  |  |  |  |
| Female | 61.54 | *ref* | — | — | — |
| Male | 68.06 | 1.33 | — | — | — |
| Marital Status |  |  |  |  |  |
| Not married | 81.48 | *ref* | — | — | — |
| Married | 64.58 | 0.41 | — | — | — |
| Field of Study |  |  |  |  |  |
| Medical Doctor | 72.88 | *ref* | *ref* | *---* | *---* |
| Pharmacy/Dentist | 38.89 | 0.24 | 0.38 | 0.15–0.98 | 0.045 |
| Nursing/Midwifery | 68.97 | 0.83 | 0.94 | 0.46–1.91 | 0.855 |
| Institution |  |  |  |  |  |
| University of Health Sciences (Vientiane) | 56.25 | *ref* | *ref* | *---* | *---* |
| Provincial institutions | 72.04 | 2.00 | 2.80 | 1.38–5.68 | 0.004 |

**Notes:**

OR = Odds Ratio; CI = Confidence Interval

Adjusted model includes age, field of study, and institution

Ethnicity and marital status were not included in the final adjusted model

*ref* = reference category

**Table 4.** Policy implications to strengthen attraction to public-sector employment among health professional students in Lao PDR

| **Policy priority** | **Core policy actions** | **Supporting evidence** |
| --- | --- | --- |
| **1. Regulate the volunteer entry pathway** | Time-bound volunteer service, basic stipends, transparent criteria for transition to contractual or civil-servant posts | Structured early-career entry systems in Thailand improved equity and retention [10,11]; aligned with national HRH reform priorities [2] |
| **2. Improve transparency of recruitment quotas** | Annual communication of national and provincial quotas, eligibility criteria and timelines | Quota transparency improved expectation–capacity alignment in Indonesia [15]; reinforced through HRH planning processes in Lao PDR [2,3] |
| **3. Strengthen career guidance during training** | Early counselling, information on public-sector entry, structured rural exposure | Early exposure and guidance are feasible and acceptable retention strategies [16]; CPIRD/ODOD experience in Thailand [11]; Workforce 2030 principles [1] |
| **4. Address central–provincial training disparities** | Targeted scholarships, incentivised rural rotations, clearer public-sector pathways | Bonded rural tracks counterbalanced urban private-sector pull in Thailand and the Philippines [9–11] |
| **5. Support early-career financial security** | Timely remuneration, housing or transport support, access to loans | Early-career support improved retention in Rwanda and Nepal [17,18] |
| **6. Integrate student intentions into HRH planning** | Routine analysis of student preferences to inform quotas and deployment | Student intention data informed workforce planning in Malaysia and similar settings [19]; aligned with Workforce 2030 and national HRH strategy [1,2] |
