## Supplementary material for "Career Intentions of Final-Year Health-Professional Students in Lao PDR: A Cross-Sectional Study of Factors Influencing Public-Sector Attraction"

**Supplementary Table S1. Additional recommendations expressed by students**

NB: These comments were collected through an open-ended survey item and are presented descriptively*.*

- To strengthen the transition of motivated graduates into the public sector, the Ministry of Health should implement a series of targeted policy reforms.

- A primary focus should be on formalizing the volunteer pathway by establishing a maximum duration for service (such as three years) and providing basic stipends to ensure early-career security.

- Recruitment must be improved through fair and transparent quota allocation, with specific priority given to long-serving rural volunteers to reward commitment to underserved areas.

- Beyond recruitment, the ministry should enhance the financial and physical well-being of staff through timely salary payments, housing support, and regular health checkups.

- Pre-service training should also be modernized to include structured career guidance and modules on professional manners and service-oriented mindsets to align student expectations with the realities of clinical practice.

- Finally, the system should adopt mechanisms to address workforce productivity, ensuring that government posts are held by active and effective personnel.
